# Types of Arrhythmias and the risk of sudden cardiac death in dialysis patients: A Systematic Review and Meta-analysis

**DOI:** 10.1101/2023.02.14.23285922

**Authors:** Subhash Chander, Sindhu Lohana, FNU Sadarat, Roopa Kumari

## Abstract

**Background:** Patients on long-term dialysis therapy due to ESRD tend to have a high mortality rate, predominantly due to cardiovascular complications, which is associated with an increased risk of arrhythmias compared to the general population. Arrhythmia has been firmly identified as the primary cause of sudden death during dialysis, as studies have shown a correlation between the timing of dialysis sessions and episodes of sudden death, as well as relationships with serum or dialysate electrotype concentrations. It also associated with a six-fold increased risk of developing ventricular fibrillation following a first myocardial infarction.

**Methods:** This systematic review and meta-analysis followed the guidelines provided by the Preferred Reporting Items for Systematic Reviews and Meta-Analyses (PRISMA). The reviewers searched the Cochrane Library, MEDLINE, Europe PMC, and Google Scholar Databases for relevant data sources. We included only randomized controlled trials and cohort studies published in English. A quantitative analysis (meta-analysis) was conducted using Review Manager version 5.4 (RevMan 5.4; The Nordic Cochrane Center, The Cochrane Collaboration, 2014).

**Results:** The initial database search yielded 547 studies, of which 213 duplicates were excluded. The title, abstract, and full-text screening excluded 247 studies, and the final total included 13 studies reporting the incidence of SCD mortality in this meta-analysis.

**Conclusion:** SCD remains a major public health concern, particularly in patients undergoing dialysis. Meta-analysis results show that bradyarrhythmia emerges as a common type of arrhythmia leading to SCD; however, other types of arrhythmias should also be considered.

## Introduction

Chronic kidney disease (CKD) affects a large population worldwide. Patients with all stages of CKD undergoing dialysis have a high prevalence of cardiovascular disease (CVD), which is associated with an increased risk of arrhythmias compared to the general population. Sudden Cardiac Death (SCD) is a major cause of mortality in such patients. Among hemodialysis patients, SCD is more common in the United States, accounting for 33% of all deaths, than in other countries such as Japan (23 %), Australia (19 %), and Canada (18 %).[1]

According to the United States Renal Data System for 2018, compared to patients with preserved kidney function, patients on maintenance dialysis have a greater risk of dying, with mortality rates exceeding 180 per 1000 patient-years.[2] Cardiovascular disease is the leading cause of death, and 40% of deaths due to a known cause are attributable to sudden cardiovascular death and arrhythmia.[3]

Sudden cardiac death (SCD) is defined as an unexpected death from cardiac causes in a person with a known or undiagnosed cardiac condition that occurs within one hour of the onset of symptoms (witnessed SCD) or within twenty-four hours of the last sign of life (unwitnessed SCD).[4] Sudden cardiac death is the primary cause of death, accounting for up to 15% of all deaths in the general population. SCD is a significant cause of death in patients with end-stage kidney disease (ESKD), but it is challenging to determine its prevalence precisely because research on SCD incidence in ESKD is frequently mixed with research on sudden cardiac arrest (SCA) that occurs during dialysis sessions.[4] On the other hand, extradialysis SCD and intradialysis SCA reflect different clinical scenarios and should be maintained. Beyond the patient’s clinical state, dialysis may favor the occurrence of life-threatening arrhythmias. Furthermore, hypotension and syncope often occur during HD sessions, and draw attention to several risk factors. To diagnose these occurrences quickly and distinguish them from SCA, healthcare personnel must immediately act upon their occurrence.[4] The basic causes of SCD are still debated, especially the precise type of terminal arrhythmia (which has important implications for prevention). Loop recorders (ILRs) used to identify terminal arrhythmias may prove useful, but a coordinated effort would be required given the low enrollment rates anticipated in such studies, according to a prescient conclusion from the Kidney Disease Improving Global Outcomes Clinical Update Conference on Cardiovascular Disease in CKD nearly five years ago. Additionally, the potential utility of Loop Recorders is not limited to the diagnosis of deadly arrhythmias. It also includes the detection of clinically undetected bradyarrhythmia and atrial fibrillation (AF). The Monitoring in Dialysis Trial (MiD), which employed ILR to identify arrhythmia in the context of HD, was the source of data on arrhythmia in dialysis patients reviewed in this publication.[5]

Although the significant risk of cardiovascular disease and sudden death is now generally acknowledged, conventional cardiovascular risk factors do not adequately explain this.[6] Particularly, there is still a lack of knowledge regarding the cause of sudden death in dialysis. Arrhythmia has been firmly identified as the primary cause of sudden death during dialysis, as studies have shown a correlation between the timing of dialysis sessions and episodes of sudden death, as well as relationships with serum or dialysate electrotype concentrations.[3]

Sudden cardiac death among hemodialysis patients and understanding its pathogenesis are quite challenging; however, some existing research studies have identified the differences between SCD among dialysis patients and normal individuals. The combination of a vulnerable myocardium and acute proarrhythmic trigger leads to terminal arrhythmias. In the general public, this condition manifests as ischemic cardiomyopathy with a reduced left ventricular ejection fraction (LVEF), which leads to disorganized cardiac conduction.[7, 8] In the early stages of chronic kidney disease, there is a high risk of sudden cardiac death, although this risk gradually increases as patients receive dialysis. The intermittent pattern of treatment exacerbates this risk by causing fluctuations in the volume and electrolyte balance.[9] Despite the fact that the risk of sudden death does not vary once dialysis is started, it is likely that the arrhythmias that lead to sudden death in dialysis patients change. It is possible that occult myocardial ischemia, intradialytic hypotension, and post-dialysis metabolic alterations, including hypokalemia, hypocalcemia, and metabolic alkalosis, contribute to the higher prevalence of ventricular tachycardia and ventricular fibrillation in the early stages of dialysis.[9] It has been shown that Patients with underlying ischemic heart disease have an increased risk of ventricular arrhythmias due to renal dysfunction.[10] It has been found that having chronic kidney disease stage 3 is associated with a six-fold increased risk of developing ventricular fibrillation following a first myocardial infarction. In implantable cardioverter-defibrillator recipients, advanced CKD has been shown to be a strong predictor of appropriate shock therapy delivery for ventricular arrhythmias. However, retrospective studies in this high-risk population did not show any survival improvement after the implantation of cardiovascular defibrillation.[9]

Although chronic kidney disease and coronary artery disease frequently coexist, sudden cardiac death is also very common in dialysis patients with no history of coronary artery disease or compromised left ventricular ejection fraction. It is possible that endothelial dysfunction, electrolyte fluxes, chronic inflammation, insulin resistance, and autonomic instability, as well as the resulting bone mineral abnormalities and vascular calcification, increase the cardiovascular risk associated with severe chronic kidney disease.[9]

Furthermore, Dalal et al. confirmed that severe ischemic cardiomyopathy and advanced chronic kidney disease increase the incidence of ventricular arrhythmias, although it remains unclear what causes SCD in dialysis patients with preserved left ventricular function.[10] Recent studies have found higher rates of bradyarrhythmic episodes and mortality in small cohorts of asymptomatic dialysis patients using implantable loop recorders (ILR). Additionally, atrial fibrillation and stroke risk are extremely high in dialysis patients.[11] Atrial fibrillation affects 20–50% of patients, according to claims data, although the exact prevalence is probably much greater because most episodes are asymptomatic. Stroke risk is 20% in patients starting dialysis, and more than half of these strokes are classified as cardioembolic or cryptogenic, with atrial fibrillation being a potential underlying cause in these cases.[9]

To optimize outcomes in patients on dialysis, it is essential to determine whether and how frequently dialysis causes potentially fatal arrhythmias. This includes identifying the types of arrhythmias that cause sudden death, particularly terminal rhythms. The need for cardiac monitoring is driven by the documentation of premature ventricular or atrial contractions, peridialytic changes in electrocardiographic morphology, and changes in heart rate variability. However, until recently, the use of surface electrocardiography (ECG) or Holter monitoring technology restricted the realistic duration of monitoring to periods that were too short to accurately capture the occurrence of arrhythmia or sudden cardiac death.[3]

Uncertainty around the “terminal event,” particularly in distinguishing sudden death from an arrhythmia from sudden death that cannot be prevented by cardiac devices, constitutes a significant knowledge gap in our understanding of sudden cardiac events in dialysis patients. The lack of autopsy data on dialysis-related SCD was identified by the Kidney Disease Improving Global Outcomes as a significant knowledge gap. It also brought attention to the limitations of traditional definitions, which include “sudden, unexpected death within an hour of symptom onset, or unwitnessed, unexpected death without obvious non-cardiac cause in patients known to be well within the past 24 hours.” In a population with a high prevalence of comorbid illness who spend a disproportionate amount of time in healthcare facilities, what exactly is “unexpected death”? Patients eventually die of terminal arrhythmias after discontinuing dialysis, although this is a withdrawal-related death. Without a patient-centered context, it would be simple to deduce incorrectly from an ILR that SCD was the “primary” event. Similarly, in the absence of rhythm tracings, conditions such as subarachnoid hemorrhage or aortic dissection may resemble SCD.[5]

It is likely complicated by certain aspects of the relationship between chronic kidney illness and arrhythmias, which remain poorly understood. In the early stages of CKD, modifiable risk factors must be identified through research initiatives. However, there is an urgent and unmet clinical need for sophisticated risk stratification strategies to identify dialysis patients who would most benefit from ICD implantation in the dialysis population where severe arrhythmic risk has already been established.[5]

### Research questions and objectives

1. To determine clinical measures to lower the likelihood of catastrophic cardiac arrest.
2. To outline how early cardiac-related illnesses can be detected
3. To elucidate the various factors involved that are linked to and predispose arrhythmias and dialysis to sudden cardiac death and elucidate potential processes and dangers
4. Increased long-term cardiac survival and a lower chance of fatal cardiac arrest in dialysis patients.
5. What are the preventive methods and options for dialysis?
6. What characteristics must be considered in the classification of rhythm abnormalities for the effective treatment of the disease?
7. What risks are generated by SCD in dialysis patients, and how do they differ from normal cardiac death?

## Methodology

### Study Design

The Preferred Reporting Items for Systematic Review and Meta-Analyses (PRISMA) guidelines were implemented using the Cochrane technique.[12] The systematic review procedure was meticulously planned to reduce bias and eliminate old subpar papers. The steps to conduct a successful systematic review are as follows: inclusion and exclusion criteria are developed in protocols, populations, interventions, comparators, and outcomes (PICO) are created, and the appropriate clinical questions are addressed. Implementation of an extensive literature review perform a search, present summaries of studies uncovered by the inquiry, and then present a few full texts.

### Literature Search

Using digital techniques, we searched numerous databases for studies. The primary databases involved in the search included PubMed, Cochrane Library, Google Scholar, Medline, and Europe PMC, and searching through the references of earlier systematic reviews and meta-analyses led to the discovery of additional papers; done to find references to helpful articles. A list of articles submitted for review and published before this comprehensive meta-analysis was conducted. Using keyword combinations and MeSH (Medical Subject Headings) phrases, a thorough computerized search was conducted to obtain results from PubMed, Medline, and Europe PMC to help create a reliable search string. The search was performed using the following keywords: Arrhythmias, Sudden Cardiac Death, Dialysis, Hemodialysis, End-Stage Renal Disease, coronary artery disease, cause of death, death, sudden cardiac arrest, SCD, coronary artery disease (cad), sudden cardiac arrest, life-threatening arrhythmia, and cardiac arrhythmia.

To obtain the references most pertinent to our search inquiry, we developed a search strategy that used Boolean operators like (AND) and (OR). Field tags (tabs and two) were also incorporated to generate variations in the search string for study identification in the databases.

### Eligibility Criteria

The reviewers evaluated the retrieved studies for eligibility by employing a pre-applied eligibility criterion. The retrieved papers were evaluated for eligibility using inclusion and exclusion criteria. Among the papers retrieved from PubMed, Medline and Euro PMC were inclusive of the following data: a population consisting of various ages, recent publications in the English language, only human participants, free, full-text papers, and study reports from years not preceding five years.

A study was considered for inclusion if it provided the intended result (risk assessment of arrhythmias, rate of sudden cardiac death in the dialysis population, and prevention of such). The following aspects were incorporated: (1) cohort studies, randomized controlled trials, clinical trials, and comparative studies comparing sudden cardiac mortality rates in dialysis and general cardiac-related illness populations. Studies considered for inclusion had to have published work on the risk factors that are prone to cardiovascular complications in dialysis patients. Findings on the development of cardiovascular complications among various age brackets of a given population should be available in the retrieved studies, (2) Single out high-risk patients undergoing dialysis or hemodialysis treatment with fatal, life-threatening cardiovascular conditions. The results of the independent eligibility assessments were combined and corrected, taking into account inputs from other parties.

### Data Extraction

The reviewers based the initial screening of data on titles and abstracts and the subsequent full-text screening of the included articles. The relevant data obtained from the selected articles were obtained using a coded Microsoft Excel sheet, which included study characteristics (author and publication year), place of study, study design, population, age, sex, arrhythmia events, SCD mortality, risk of SCD, and conclusion. Study quality was assessed and judged using the Review Manager version 5.4 (RevMan 5.4: The Nordic Cochrane Center, The Cochrane Collaboration, 2014).

## Results

### Study Selection

The initial search generated 5816 studies. A total of 1960 studies were excluded for duplication reasons, while 3856 articles were included in the title and abstract screening, where 3326 studies were excluded for eligibility reasons. Next, the reviewers performed full-text screening of 530 studies to eliminate 519 additional studies. Finally, 11 studies were included in the analysis, and two more studies were selected from the bibliography list of previous reviews, bringing the final tally to 13 articles that were used for data extraction. Figure 1 shows the PRISMA 2020 flow diagram for updated systematic reviews, summarizing the selection process.

**Figure 1:**
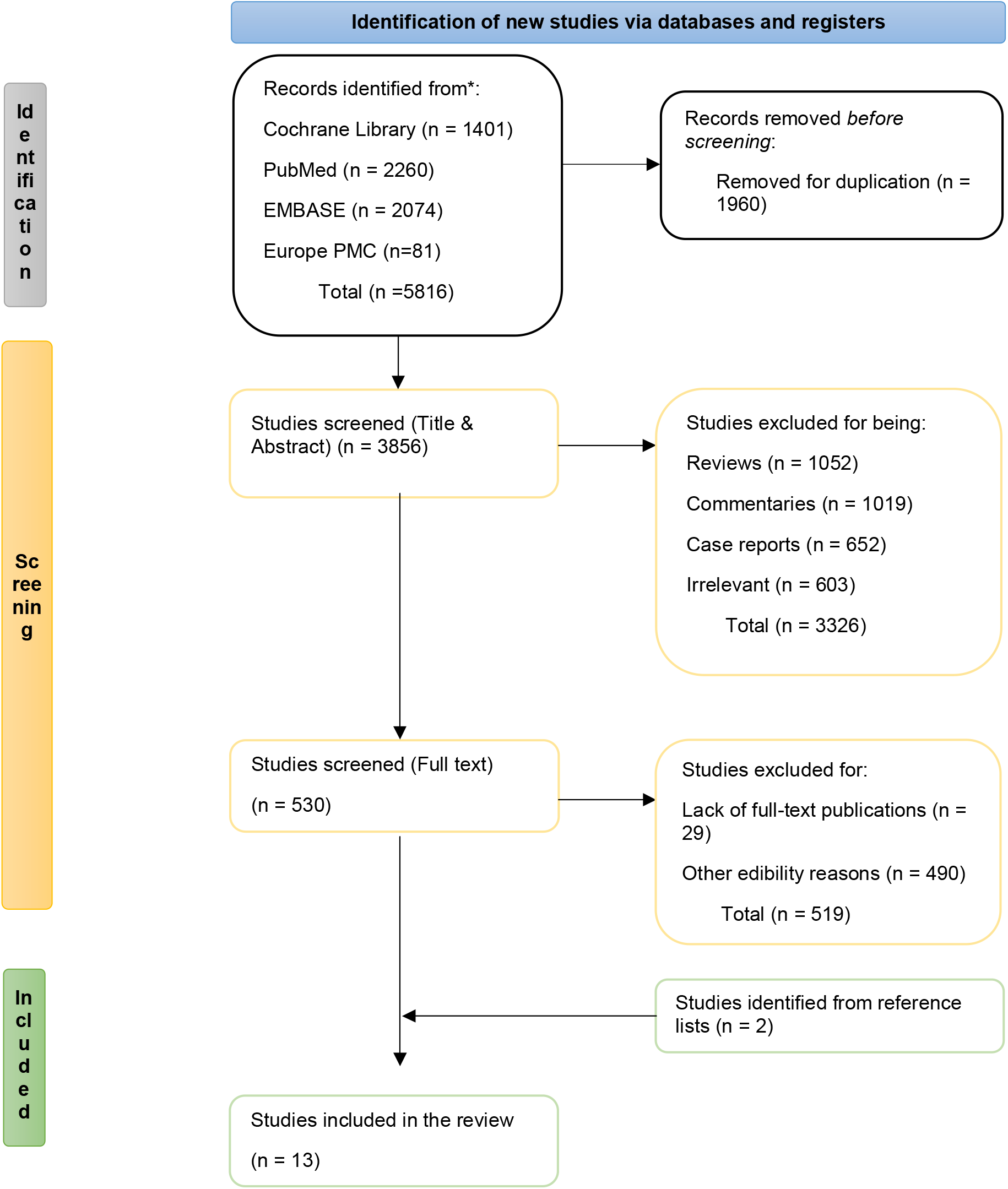
PRISMA flow diagram for the study selection for the systematic review.

| Study Characteristics |  | Study Design | Population | Age | Gender (M/F) | Arrhythmia events | SCD Mortality | Risk of SCD (HR; 95% CI) | Conclusive Findings |
| --- | --- | --- | --- | --- | --- | --- | --- | --- | --- |
| Author | Year |  |  |  |  |  |  |  |  |
| Pfau et al.<sup>[13]</sup> | 2021 | RCT | 1108 | 65.6 (8.5) years - 67.6 (8.1) years | 603/505 |  | 139 | 1.62; 1.03 to 2.56 | In individuals receiving dialysis, elevated serum oxalate represents a new risk factor for cardiovascular problems and sudden cardiac death. |
| Jukema et al.<sup>[14]</sup> | 2019 | RCT | 188 | ≥55 years - <81 years | 188 Males |  | 19 | 1.32; 0.53 to 3.29; | Prophylactic ICD therapy did not lower the rate of SCD or all-cause mortality, which remained high, in a cohort that was well screened and given appropriate care while receiving dialysis. |
| Tangri et al.<sup>[15]</sup> | 2011 | RCT | 1747 | 58 years | 751/996 |  | 181 | 0.65; 0.42 to 1.01 (with IHD) 1.61; 0.92 to 2.80 (without IHD) | Use of β-blockers was not linked to a decreased incidence of sudden cardiac mortality in hemodialysis patients without a history of IHD. |
| Cheung et al.<sup>[16]</sup> | 2004 | Randomized Multi-Center Trial | 1846 | 18 - 80 years | 808/1038 |  | 343 | 2.57; 1.73 to 3.83 | According to the HEMO Study, ischemic heart disease (IHD) is a significant contributor to cardiac hospitalizations and cardiac deaths. |
| Karnik et al.<sup>[17]</sup> | 2001 |  | 400 | Mean Age 66.3 years (≥75 years and ≥85 years) | 206/194 | atrial fibrillation, atrial flutter, supraventricular tachycardia, bradycardia, sick sinus syndrome, first degree heart block, trifascicular block, unspecified heart block, left bundle branch block, right bundle branch block, unspecified ventricular arrhythmia, ventricular tachycardia, ventricular fibrillation, and asystole. | 14 |  | A relatively rare yet serious side effect of hemodialysis is cardiac arrest. Dialysate prescriptions for high-risk patients should be continuously reviewed and changed in order to lower the risk of adverse cardiac events while undergoing hemodialysis. |
| Wang et al.[18] | 2010 | Cohort Study | 230 |  | 117/113 |  | 28 | 78.6% (95% CI: 72.2% to 84.1%) | To identify patients at risk of sudden cardiac death, N-terminal probrain natriuretic peptide may be utilized instead of echocardiography, although it offers no additional benefit in terms of predicting sudden cardiac death. |
| Wanner et al.[19] | 2005 | Clinical Trial; multicenter, randomized, double-blind, prospective study | 1255 | 18 years to 80 years | 1255 Females |  | 160 | 0.81 (0.64–1.03) | In patients with diabetes receiving hemodialysis, atorvastatin had no statistically significant impact on the composite primary end point of cardiovascular death, nonfatal myocardial infarction, and stroke. |
| Drechsler et al.[20] | 2011 | RCT | 1255 | 18 years to 80 years | 671/584 |  | 160 | 3.1; 2.0-4.9 and 2.4; 1.5-3.9 | Low homoarginine levels in dialysis patients are a significant risk factor for SCD and heart failure-related mortality. |
| Kerns et al.[21] | 2018 | Cohort Study | 558 | 18 years and above (Mean Age 56 years) | 312/246 |  | 30 | 3.62; 1.36-9.66 | Patients receiving incident hemodialysis who have Obstructive sleep apnea (OSA) are more likely to die from all causes, have cardiovascular disease, and develop SCD. |
| silva et al.[22] | 2015 | Cohort Study | 100 | Mean Age, 59 ± 8, 8 years | 65/35 | Bradyarrhythmias, Asystole, Bradycardia, Advanced atrioventricular block, Premature atrial beats, Sinusal tachycardia, non-sustained atrial tachycardia, Sustained atrial tachycardia, Atrial fibrillation, Premature ventricular beats, Non-sustained ventricular tachycardia, Sustained ventricular tachycardia/ventricular fibrillation, Supraventricular (94%) and ventricular arrhythmias (79%) | 7 | 2.83; 95% CI, 1.01-7.96 | Implantable loop recorders (ILR) helped identify a significant prevalence of arrhythmic events (AE), the majority of which lacked clinical value, in the medium-term follow-up of renal transplant candidates (RTCs). |
| Sacher et al.[23] | 2018 | RCT | 71 | Mean Age 65 ± 9 years | 52/21 | Bradycardia, asystole, ventricular arrhythmia | 4 | 46.23; 7.96–268.48 | To enable early detection and the start of appropriate treatment, ILR may be taken into consideration in HD patients who are at risk for serious conduction abnormalities, ventricular arrhythmia, or atrial fibrillation or flutter. |
| Wheeler et al.[24] | 2014 | Randomized, Double-Blind, Placebo-Controlled Clinical Trial | 3883 | ≥18 years | 3,883 |  | 348 | 0.79; 0.64 to 0.98 | Accepting the limits of post hoc analysis, any advantages of cinacalcet for the treatment of cardiovascular disease in the context of hemodialysis may come from the reduction of nonatherosclerotic processes. |
| Roberts et al.<sup>[25]</sup> | 2017 | Cohort Study | 30 | 68±12 years | 18/12 | Bradyarrhythmia and tachyarrhythmia | 2 | 0.87; 0.66–1.13 | Contrary to original assumptions, bradyarrhythmias appeared as a frequent and possibly serious arrhythmic event, confirming the high death rate observed in hemodialysis populations. |

#### Meta-Analysis

Thirteen studies that reported the incidence of SCD mortality were included in this meta-analysis. The summary statistics of the dataset generated a standard deviation of 124.4799 (Figure 3). A meta-analysis was then performed, outputting 110.38 [42,72, 178.05] effect size at a 95% confidence interval. There was a low level of heterogeneity (I-squared = 0.00%) and a p-value of 0.00, which was less than the significance level of 0.05. The present meta-analysis proved the significance of SCD mortality among dialysis patients. Figures 4 and 5 show the forest and funnel plots representing the outcome of the meta-analysis.

**Figure 3:**
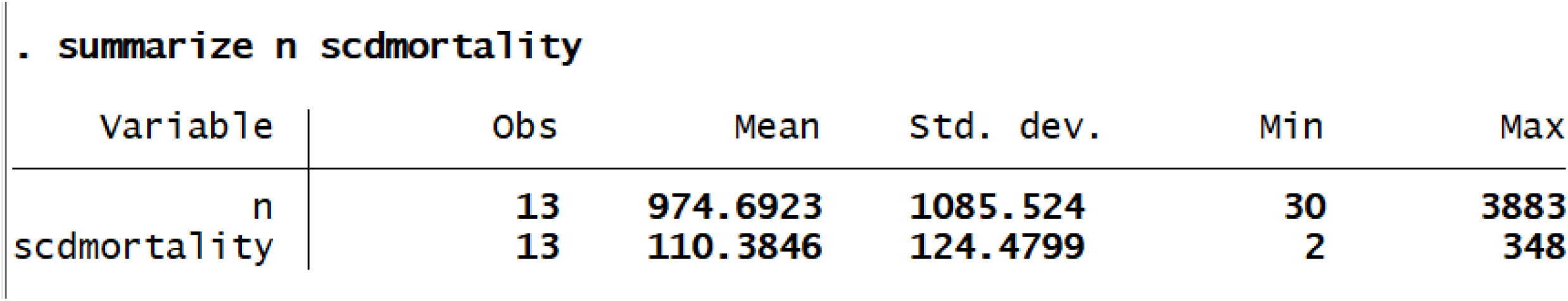
Summary statistics of the dataset calculating the standard deviation.

**Figure 4:**
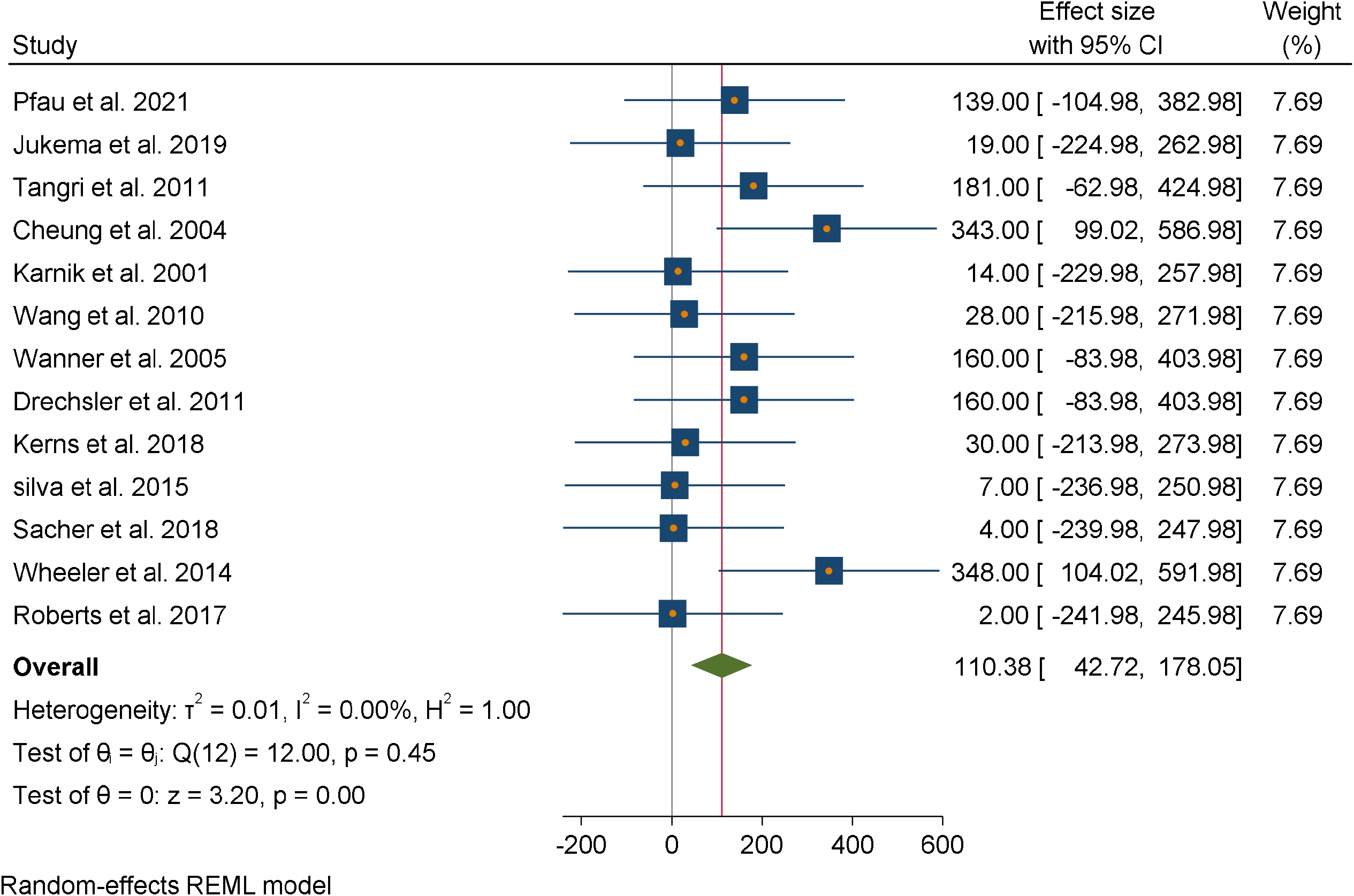
A forest plot for the meta-analysis of the incidence of SCD mortality in dialysis patients.

**Figure 5:**
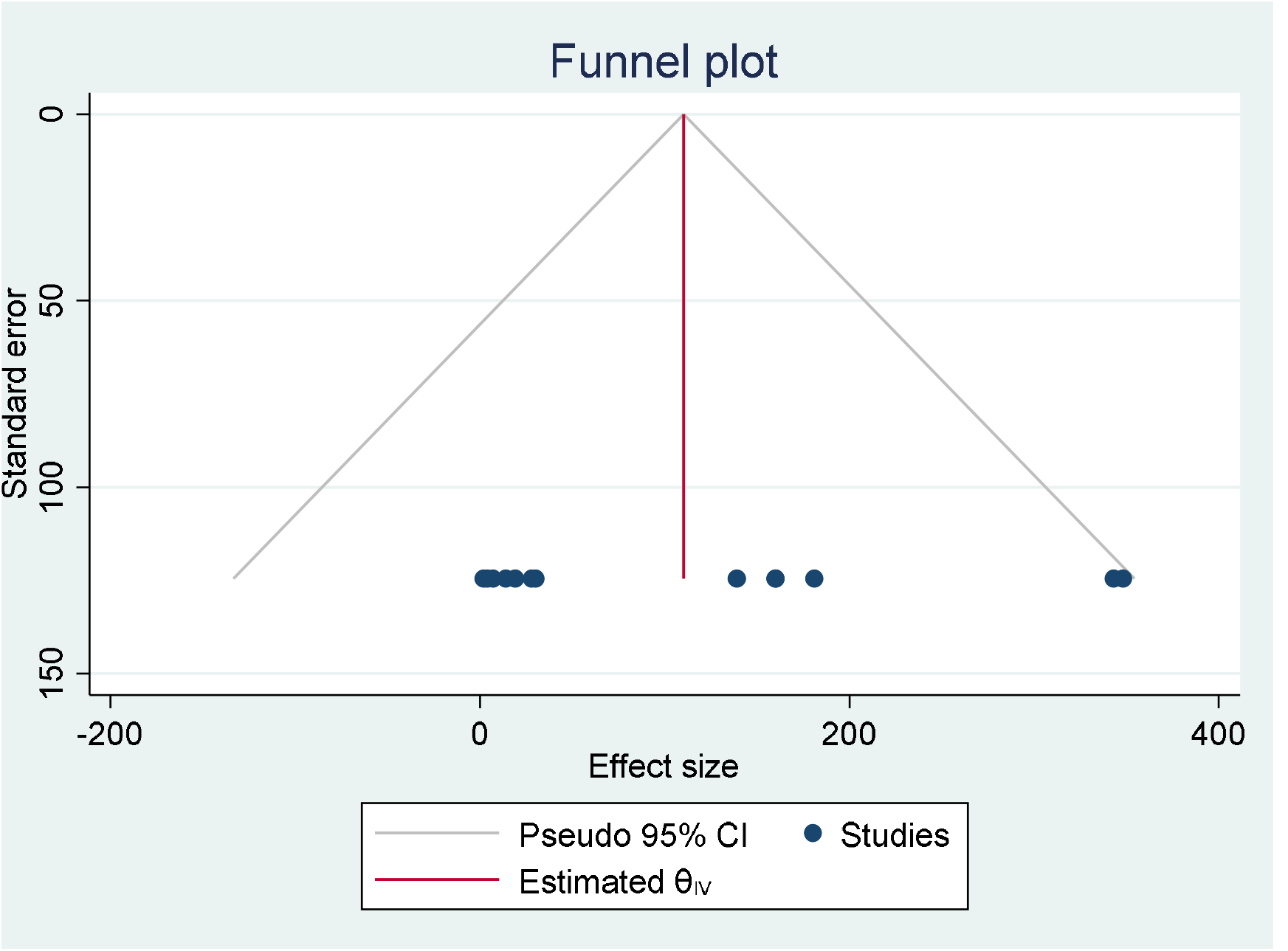
Funnel plot representing publication bias between the analyzed studies.

## Discussion

Our meta-analysis analyzed cohort studies and randomized controlled trials that investigated the types of arrhythmias and the risks of cardiac death among dialysis patients. Patients on long-term dialysis therapy due to ESRD tend to have a high mortality rate, predominantly due to cardiovascular complications.[21, 22, 24] Meta-analysis results showed that SCD commonly occurs in patients undergoing dialysis. These findings suggest a primary increase in the risk of arrhythmias (the leading cause of SCD).[23] This group of patients has several predisposing factors that put them at a high risk of arrhythmia. Multiple studies have shown that arrhythmias occur in all age groups; however, the fatality rate is higher in dialysis patients due to cardiovascular complications. Therefore, the meta-analysis focused on the risk assessment of different types of arrhythmias to evaluate the mortality rate resulting from cardiac deaths in dialysis patients. Multiple studies have proposed the factors that increase the risk of cardiac arrest, complications, and death, including inflammation, hypertension, hypercoagulability, and autonomic dysfunction.[16, 19, 20]

### Sudden Cardiac Death in Dialysis Patients

Roberts et al. monitored arrhythmia and SCD in dialysis patients and observed that in 379 512 hours, there were eight deaths. The study reported a 95% confidence interval for median event-free survival for any type of arrhythmia. These findings confirm that a high mortality rate is recorded in the dialysis population, which requires further research for better management. The prevalence and severity of cardiac arrest risk factors and stressors in dialysis patients are unique to dialysis sessions and result in half of all hemodialysis-related deaths caused by cardiovascular complications. Sacher et al. also indicated that cardiac arrest accounted for nearly one-third of all cardiac mortality rates in this group of patients.[23] A case-cohort study by Wang et al. illustrated that patients who experienced inpatient cardiac arrest were older and had diabetes.[18] However, multiple studies have not established any association between race or gender and SCD, although men seem to have an increased risk of ischemic heart disease compared to women. Approximately half of these patients had a medical history of coronary artery disease, while 1 in 6 patients had a significant systolic blood pressure drop (≥ 30 mmHg) before cardiac arrest. Our meta-analysis illustrated systolic dysfunction as the most significant predictor of SCD, followed by either low diastolic or high systolic blood pressure. According to Karnik et al., the results agree with data from the general population, indicating that reduced function in the ventricle mainly predicts the presence of SCD.[17] Jukema et al. showed that poor systolic function predisposes patients to heart failure, which can increase the risk of ventricular arrhythmia through neurohumoral activation.[14]

### Classification of Rhythm Abnormalities

Bradyarrhythmia and tachyarrhythmia are the most common rhythm abnormalities causing cardiac arrest in a dialysis patient. Recent studies on Implantable Loop Recorders (ILR) have illustrated that bradyarrhythmia significantly contributes to sudden death in these patients. However, most of them fail to include the incidence of dialysis patients with high cardiovascular complications that are at a greater risk of tachyarrhythmias. The analysis results suggest that fatal or serious arrhythmias in dialysis patients represent defects such as asystole or bradycardia, rather than tachyarrhythmias. These outcomes are echoed by a clinical study by Tangri et al. (2011), who reported that an episode of a paused heart or bradycardia mostly precedes 50% of cardiac deaths.[15] Furthermore, Pfau et al. (2021) emphasized this hypothesis with the findings that the majority of patients received a pacemaker or a defibrillating device following recommendations to do so to improve their conditions.[13] However, all patients experience arrhythmia as the final event, which may be either VF or asystole before death occurs. Such cases were observed and reported in approximately ten people who had progressive asystole or bradycardia. Data has suggested that there should be further clinical studies on devices with monitored and combined defibrillation and pacing capacity owing to the lack of sufficient information on the efficacy of ICDs.

### Management of SCD in Dialysis Patients

According to Pfau et al., Cheung et al., and Drechsler et al., a common-sense approach to preventing Sudden Cardiac Death among dialysis patients would be first identifying and then treating the population who are most vulnerable using the existing cardiovascular medications and reducing potential triggers to arrhythmic events.[13, 16, 20]. Tangri et al. (2011) pointed out that strategies to minimize such triggers entail measures such as extended or more frequent dialysis sessions, avoiding potassium consumption, reducing dialysate temperatures, and monitoring potassium levels prior to dialysis sessions.[15] Further, a study by Wanner et al. recommends other therapies to manage SCD, including the use of appropriate therapy, that includes anti-arthymic pharmacological agents, implantable cardiac defibrillators (ICD), and pacemakers.[19] However, these devices should be applied on a case-by-case basis because of the existing hazards that they might present to dialysis patients.

#### Strength and Limitations

A major strength of our meta-analysis concerning the prevalence of SCD is that trained and reputable reviewers carefully conducted data extraction to produce high-quality information on the study topic. However, the analyzed clinical studies on the incidence SCD due to arrhythmia in dialysis patients vary widely, while studies focusing only on adults or older people are scarce. Another limitation arises during multiple statistical comparisons of influences between randomized controlled studies; thus, interpretation of the additional analysis outcomes should be cautiously performed to avoid data misinterpretation.

### Study Implications and Future Perspectives

Our meta-analysis emphasizes the need for further studies to determine the incidence of SCD in dialysis patients, which remains a major health concern. These clinical studies should focus on developing risk stratification strategies for bradyarrhythmia and tachyarrhythmia. In particular, it is important to identify the specific type of arrhythmic event that may cause Sudden Cardiac Death in dialysis patients; thus, despite recent advances, it is essential to have future studies focused on this area of study.

## Conclusion

SCD remains a major public health concern, particularly in patients undergoing dialysis, as there is a high risk of cardiac arrest, which has the highest mortality rate among this group. Bradyarrhythmia emerges as a common type of arrhythmia leading to SCD; however, other types should also be considered. Owing to the potential to improve cardiac outcomes, additional strategies should be developed to minimize cardiac mortality and morbidity in managing the dialysis population. In addition, it is essential to note the significance of the early detection and treatment of cardiovascular complications to reduce fatality rates.

## Data Availability

All data produced in the present work are contained in the manuscript

## Statement of Ethics

An ethics statement is not applicable because this study is based exclusively on published literature.

## Author Contributions

In the Author Contributions section, a short statement detailing the contributions of each person named as an author should be included. Contributors to the paper who do not fulfil the <u>ICMJE Criteria for Authorship</u> should be credited in the Acknowledgement section.

**Subhash Chander:** Created the hypothesis, literature review and prepare the discussion part and analysis.

**Sindhu Lohana:** Literature search and prepare the introduction part. FNU Sadarat: Extract the data, formulate the tables and graphs.

**Roopa Kumari:** Analyze the data, results, screened the studies and finalize the draft.

## Data Availability Statement

Data can be provided and available on request.

